# Effective deep brain stimulation for obsessive-compulsive disorder and Tourette Syndrome increases network-wide neural variability

**DOI:** 10.1101/2025.10.09.25337425

**Authors:** Timon Merk, Kalman A. Katlowitz, Tommy Liu, Sandesh Reddy, Anthony Allam, Jonathan H. Bentley, Kasra Mansourian, Sarah Soubra, Thomas Hamre, Rick R. Hanish, Garrett P. Banks, Arjun Tarakad, Steven T. Bellows, Nora Vanegas-Arroyave, Eric A. Storch, Sarah R. Heilbronner, Wayne K. Goodman, Sameer A. Sheth, Nicole R. Provenza

## Abstract

**Introduction:** Deep brain stimulation (DBS) is an effective therapy for obsessive-compulsive disorder (OCD) and Tourette syndrome. Our previous results demonstrated disrupted circadian periodicity and increased neural variability in the region of the ventral striatum (VS) in responders to DBS for OCD. Here, we tested the hypothesis that the relationship between increased neural variability and symptom improvement is not specific to the ventral striatum alone but generalizes to another basal ganglia region, the globus pallidus internus (GPi).

**Materials and Methods:** We gathered chronic local field potential (LFP) recordings from four patients with OCD and comorbid Tourette syndrome implanted with bilateral dual-site DBS leads in the ventral striatum VS and GPi. We measured spectral power in the 9 Hz band in both targets continuously for over 900 days using the DBS system’s on-device recording capability. We estimated neural variability using an autoregressive model applied to the continuous time series of neural power.

**Results:** Two of the four patients achieved responder status for both OCD and TS. In all four patients, neural variability accurately classified responder status from LFP recordings in both VS (for OCD) and GPi (for TS) targets. Cross-correlation and Granger causal analyses between the two targets showed different lead-lag intervals and directionality, indicating non-redundant information content.

**Conclusion:** Our findings demonstrate that increased low-frequency neural variability within the VS and GPi predicts symptom improvement in OCD and TS, respectively. These findings extend our previous results in OCD alone and recapitulate the notion that biological systems function optimally with a healthy degree of physiological variability.

## Introduction

Deep brain stimulation (DBS) has emerged as an effective neuromodulatory therapy for treatment-resistant neuropsychiatric disorders, including obsessive-compulsive disorder (OCD) (Gadot et al., 2022) and Tourette syndrome (TS) (Martinez-Ramirez et al., 2018). Despite growing clinical success, the underlying mechanisms by which DBS exerts its therapeutic effects remain poorly understood. Increasing evidence suggests that, in contrast to movement disorders such as essential tremor or Parkinson’s disease, these effects are neither immediate nor local and involve complex alterations in circuit-level and temporal dynamics of brain activity (Bourne et al., 2012). One such component is the role of circadian rhythms in shaping neural excitability, neurotransmission, and symptom expression (Nota et al., 2015; van Rheede et al., 2022). Yet, circadian patterns of subcortical neurophysiology in patients with neuropsychiatric disorders remain largely unexplored.

Recent advances in implantable sensing-enabled neurostimulators, such as the Medtronic Percept, allow for chronic longitudinal recording of local field potentials (LFPs) in patients with these DBS system (Stanslaski et al., 2024). This technology provides an opportunity to investigate how naturally occurring and potentially pathological neural rhythms interact with disease states and neuromodulation in real-world conditions. We previously demonstrated that low neural variability predicted clinical response in OCD patients in the region of the ventral striatum (VS) (Provenza et al., 2024). A remaining open question is whether this outcome-predicting pattern is unique to the DBS target region or is also evident in other regions of the network. In this study, we made use of the unique opportunity to investigate the nature of chronic circadian neural patterns in two different subcortical regions within the same patients across months of recordings at home. We implanted DBS electrodes in the VS and globus pallidus internus (GPi) to treat comorbid OCD and TS, respectively (Najera et al., 2023). This rare clinical setup allowed us to examine LFP recordings from two anatomically and functionally distinct targets. Since both targets are used independently to treat different neurological disorders (Provenza et al., 2024; Shahed et al., 2007; Shofty et al., 2023), and they have unique roles in the intricate cortico-basal ganglia circuitry central to these disorders, we hypothesized that there would be independent DBS-modulated neural rhythmicity. In this cohort we tested whether this neural biomarker also tracks symptom improvement in a different subcortical region and a different neuropsychiatric disorder.

Through time-resolved spectral analysis and autoregressive modeling of LFP power in the 9□Hz range, we show that circadian rhythmicity was present in both GPi and VS regions prior to stimulation. Surprisingly, both became disrupted in responder patients following therapeutic DBS. We therefore explored the relationships between the two regions and found that GPi and VS rhythms were correlated for a subset of patients without a temporal delay. In other patients, coupling with different time-lags across several hours was present for both responder and non-responder patients. For an exemplary patient we show that the 9 Hz coupling was not present within short high-temporal resolution recordings, but coupling became evident within multiple-hour longitudinal measurements. These findings provide new insights into the chronobiology of subcortical circuits and highlight the potential of circadian biomarkers of subcortical multi-target approaches for guiding and assessing neuromodulatory therapies.

## Methods

### Participants

Four adults with a primary diagnosis of severe, treatment-resistant obsessive-compulsive disorder (OCD) comorbid with Tourette syndrome (TS) underwent bilateral deep brain stimulation (DBS) implantation following informed consent. The study protocol was approved by the Institutional Review Board (IRB) at Baylor College of Medicine (H-49155). Gender (as presented in Table 1) was self-reported. Sex and gender were not considered in the study design, and no sex/gender-based analyses were conducted due to the small sample size. Participants were not financially compensated for their involvement in the study. All patients were monitored before and after DBS activation. Stimulation onset time points were separate for GPi and VS stimulation. Detailed patient characteristics are shown in Table 1. Detailed descriptions of case selection, surgical protocols, and core data analysis have been previously reported (Najera et al., 2023; Provenza et al., 2024; Shofty et al., 2023).

**Table 1:** Patient demographics, clinical outcomes and duration of continuous neural recordings. Gender was determined based on self-report. Follow-up duration was calculated as the time between initial surgery and the most recent clinical visit. OCD responder status is defined as ≥35% reduction in the Y-BOCS II. TS responder status is defined as ≥50% reduction in the YGTSS

| Patient | Gender | Age Range (years) | Follow-up duration (months) | OCD Responder Status | Initial Y-BOCS II score | Final Y-BOCS II score | Percent Y-BOCS II reduction | TS Responder Status | Initial YGTSS score | Final YGTSS score | Percent YGTSS reduction | VS LFP recording duration days (L\|R) | GPi LFP recording duration days (L\|R) |
| --- | --- | --- | --- | --- | --- | --- | --- | --- | --- | --- | --- | --- | --- |
| P001 | F | 41-45 | 35 | Yes | 41 | 26 | 36,59% | Yes | 80 | 17 | 78,75 % | (150\|150) | (150\|150) |
| P005 | M | 16-20 | 20 | Yes | 42 | 14 | 66,67% | Yes | 94 | 36 | 61,70 % | (319\|319) | (293\|293) |
| P008 | F | 31-35 | 23 | No | 36 | 26 | 27,78% | No | 76 | 49 | 35,52 % | (260\|239) | (251\|251) |
| P010 | F | 21-25 | 9 | No | 32 | 28 | 12,50% | No | 65 | 62 | 4,62% | (190\|190) | (101\|102) |

### Surgical implantation

DBS leads (Medtronic model 3387 or SenSight, 1.5-mm contact spacing) were implanted bilaterally in the globus pallidus internus (GPi) and ventral capsule/ventral striatum (VC/VS), with electrode placement confirmed by intraoperative imaging and awake physiological testing. Each lead pair was connected to extensions tunneled to independent bilateral Medtronic Percept PC (P1) or RC (P2, P3, P4) pulse generators. Clinical status was assessed using the Yale-Brown Obsessive-Compulsive Scale (Y-BOCS; (Storch et al., 2010)) for OCD and the Yale Global Tic Severity Scale (YGTSS; (Leckman et al., 1989)) for TS. These assessments were conducted preoperatively and at regular intervals throughout the course of DBS therapy. OCD clinical response was defined as a ≥35% reduction in Y-BOCS score or, qualitatively, as a substantial improvement enabling return to professional or social functioning. Time periods labeled as “unlabeled” reflect either indeterminate clinical status (due to insufficient data) or intermediate improvement (i.e., not meeting full response criteria).

### Invasive electrophysiological recordings

We acquired time-domain local field potential (LFP) recordings directly from the Medtronic Percept PC or RC neurostimulators at a sampling rate of 250□Hz during scheduled clinic visits. In addition to raw time-domain data, the Medtronic Percept enables chronic sensing by providing a continuous estimate of the average LFP amplitude within a user-specified frequency band (center frequency ±2.5cHz). This power spectral data was sampled and stored onboard the device every 10 minutes, yielding 144 data points per day. We retrieved data using the Medtronic DBS Clinician Programmer during follow-up visits. For chronic sensing, we configured the device to monitor the LFP amplitude (expressed in microvolts peak) in the 8.79□±□2.5□Hz frequency band, which we refer to as “9□Hz band” throughout the manuscript. We exported these long-term data in JSON format during clinical sessions for further analysis and converted timestamps from Coordinated Universal Time (UTC) to the local time zone. We z-score normalized daily power values using respective daily means and standard deviations to account for gradual increases in baseline LFP amplitude and stimulation intensity over time.

### Computation of circadian neural periodicity

We generated circular polar plots for each day to visualize daily fluctuations in 9-Hz power across the 24-hour cycle. To assess the periodicity and temporal structure of the signals, we applied two model-based characterizations: the coefficient of determination from cosinor analysis (cosinor R^2^) and separately linear autoregressive R^2^. We computed state-level averages of autoregressive R^2^ for the pre-DBS severe OCD condition and the post-DBS clinical response condition (Figure 3c). We calculated daily estimates of both metrics to generate distributions representing each clinical state and their respective averages. In the following sections, we describe each metric used to quantify neural circadian periodicity. For cosinor estimates we fitted the normalized LFP data to a sum of cosine functions using a linear regression model, described by Moškon et al. 2020 (Moškon, 2020) implemented via the statsmodels package (v0.14.0). For each analysis day, we applied the cosine model within a two-day window centered on the day of interest. From these fits, we extracted the acrophase (the timing of the daily cosine peak relative to 00:00 local time), amplitude, and coefficient of determination (R^2^). We plotted the resulting phase and amplitude values for each patient as polar plots, using only the primary (i.e., largest) peak to define the reported acrophase. To evaluate circadian rhythmicity across clinical states, we fitted the cosinor model separately to each state—pre-DBS, persistent OCD symptoms, and clinical response— using a five-fold cross-validation strategy applied to all 10-minute timepoints. For each fold, we predicted the z-score normalized LFP signal and calculated the corresponding R^2^ to quantify model performance. We averaged the R^2^ values across folds to generate a single cross-validated R^2^ per clinical state for each patient. Additionally, we aggregated all predicted timepoints to compute a daily R^2^ value, reflecting the model’s explanatory power on a day-by-day basis. We performed all R^2^ computations using the scikit-learn package (v1.2.2) (Pedregosa et al., 2011) and expressed the final R^2^ values as follows:

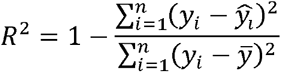

where *y*_*i*_ is the true value; is the predicted value at sample *i*; and 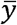 is the mean of true values.

#### Linear autoregressive model analysis and statistics

A linear autoregressive (AR) model predicts time-series data by expressing the current value *X*_*t*_ as a linear combination of its previous values, along with a constant term and a noise component. The model takes the form:

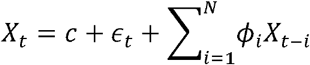

To compare neural dynamics between clinical states, we calculated the mean difference of the linear autoregressive R^2^ between the pre-DBS and post-DBS response states. We used this delta to quantify how these metrics changed with symptom improvement. To assess the separability of symptom states, we trained a maximum margin classifier. This classifier defined a linear decision boundary separating the average delta values and average daily output measures across clinical states. We then compared the linear autoregressive R^2^ values between pre-DBS and post-DBS distributions within each patient using two-sample, two-tailed Welch’s t-tests.

#### Analysis of longitudinal recordings

We performed different analysis to quantify the coupling of longitudinal and high-temporal recording oscillations between the two targets. First, we computed 2d-histograms of instantaneous intra-day z-score normalized power distributions. We fitted a linear regression function between the GPi and VS power values (linear regression scikit-learn v1.6.1) (Pedregosa et al., 2011) and computed the Pearson correlation coefficient between both longitudinal power series. Next, we computed the cross-correlation function for each day and estimated the maximum value without the zero time-lag component. We computed the separate mean cross-correlation (scipy v1.15.2) (Virtanen et al., 2020) for each responder patient during clinical stimulation response and non-response periods. Furthermore, we tested if the response- and non-response mean cross-correlation patterns were statistically different by computing permutation tests (Monte Carlo n=5000 permutations). Additionally, we computed Granger causality (statsmodels v0.14.4) (Seabold and Perktold, 2010) with a maximum lag of 12 hours on 30 min group-averaged data. We estimated F-test statistics on linear regression models using the sum of squared residuals. We evaluated the scores in VS to GPi and GPi to VS direction separately, and the net score as the difference of VS->GPi and GPi->VS scores.

## Results

Four patients (3F, mean age 29 ± 10.5 years, Table 1) with comorbid OCD and Tourette Syndrome (TS) met surgical criteria to be treated with bilateral DBS in the ventral capsule/ventral striatum (VC/VS) and globus pallidus internus (GPi) (Figure 1a). Each pair of leads was connected to its respective implanted pulse generator, allowing for chronic stimulation and sensing of neural activity in both brain regions. Reproducing previous findings (Provenza et al., 2024), we identified a narrow-band spectral peak at the theta/alpha border (~9 Hz) in the VS region in every patient (Figure 1b, orange). In the GPi region the same theta/alpha border peak was present within each patient as well (Figure 1b, green). We recorded average power around this spectral peak in 10-minute increments with a mean follow-up time of 662 ± 325 days. Throughout this lengthy period we assessed severity for both OCD and TS symptoms using standard clinical assessments (Figure 1c). The stimulation response criterion was defined as a 35% reduction in symptoms on the Y-BOCS-II (Storch et al., 2010) for OCD, and separately a 35% reduction in symptoms on the YGTSS (Leckman et al., 1989) for TS. Two patients responded to stimulation both for OCD symptoms (YBOCS-II pre-OP→post-OP P001: 41→26, P005: 42→14) and Tourette’s symptoms (YGTSS pre-OP→post-OP P001 80→17, P005: 94→36). The other two patients were classified as non-responders for OCD (YBOCS-II pre-OP→post-OP P008: 36→26, P010: 32→28) and Tourette’s symptoms (YGTSS pre-OP→post-OP P008: 76→65, P010=65→62). GPi stimulation and VS stimulation were activated at different time points, with GPi stimulation preceding VS stimulation, as guided by clinical factors.

**Figure 1.**
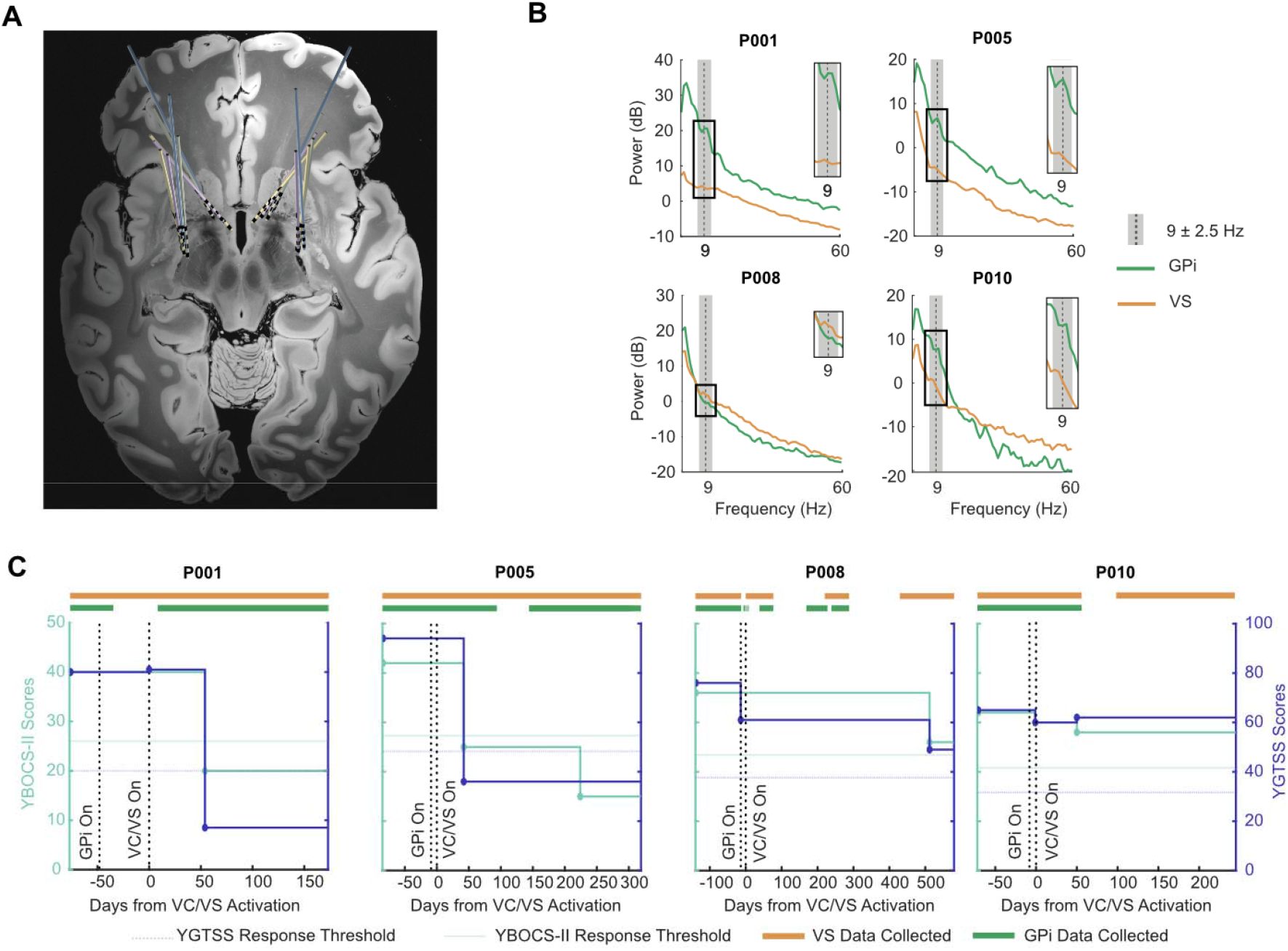
Concurrent GPi and VS stimulation for OCD and comorbid Tourette syndrome. a) Axial MRI showing bilateral deep brain stimulation (DBS) lead placements in GPi and VS targets for all patients (n=4). b) Exemplar GPi and VS power spectral density plots show 9 ± 2.5 Hz range peaks in the VS. All recordings from left lead. c) Longitudinal clinical response. Y-axes show scores from the Yale-Brown Obsessive Compulsive Scale-II (YBOCS-II, left) and Yale Global Tic Severity Scale (YGTSS, right). Days aligned to VS stimulation activation (day 0 marked by vertical dashed line) are shown in addition to GPi stimulation activation. Light horizontal lines mark the minimum 35 % symptom reduction for each patient and disease condition.

### Disruption of neural rhythmicity in both GPi and VS classifies clinical stimulation response

We recorded 9 ±□2.5□Hz power in both targets across both hemispheres using chronic passive recordings at 10 min intervals. Given our previous results indicating that left hemisphere VS recordings are more predictive of clinical response than that of the right hemisphere, we limited our analyses to left hemisphere recordings only (Provenza et al., 2024). Because stimulation was initiated weeks after DBS implantation, we had access to recordings not only after stimulation initiation but also before. On average we recorded 34.75 ± 8.79 days before and 457.25 ± 297.6 days after GPi stimulation initiation; and 54.5 ± 11.59 days before and 430.75 ± 314.88 days after VS stimulation initiation. We used linear autoregressive (AR) modelling with a time lag of one day (10 minute samples, total 144 samples) to predict spectral power dynamics. We first reproduced previous findings that the 9 Hz circadian pattern in the VS showed clear disruption aligned to OCD clinical response within VS (Figure 2b) (Provenza et al., 2024). We now report that this disruption is not isolated to the VS, but also extends to the GPi (Figure 2a), and is correlated with TS clinical response. In both non-responder patients, the neural circadian rhythmicity remained stable throughout the recording duration in both targets.

**Figure 2.**
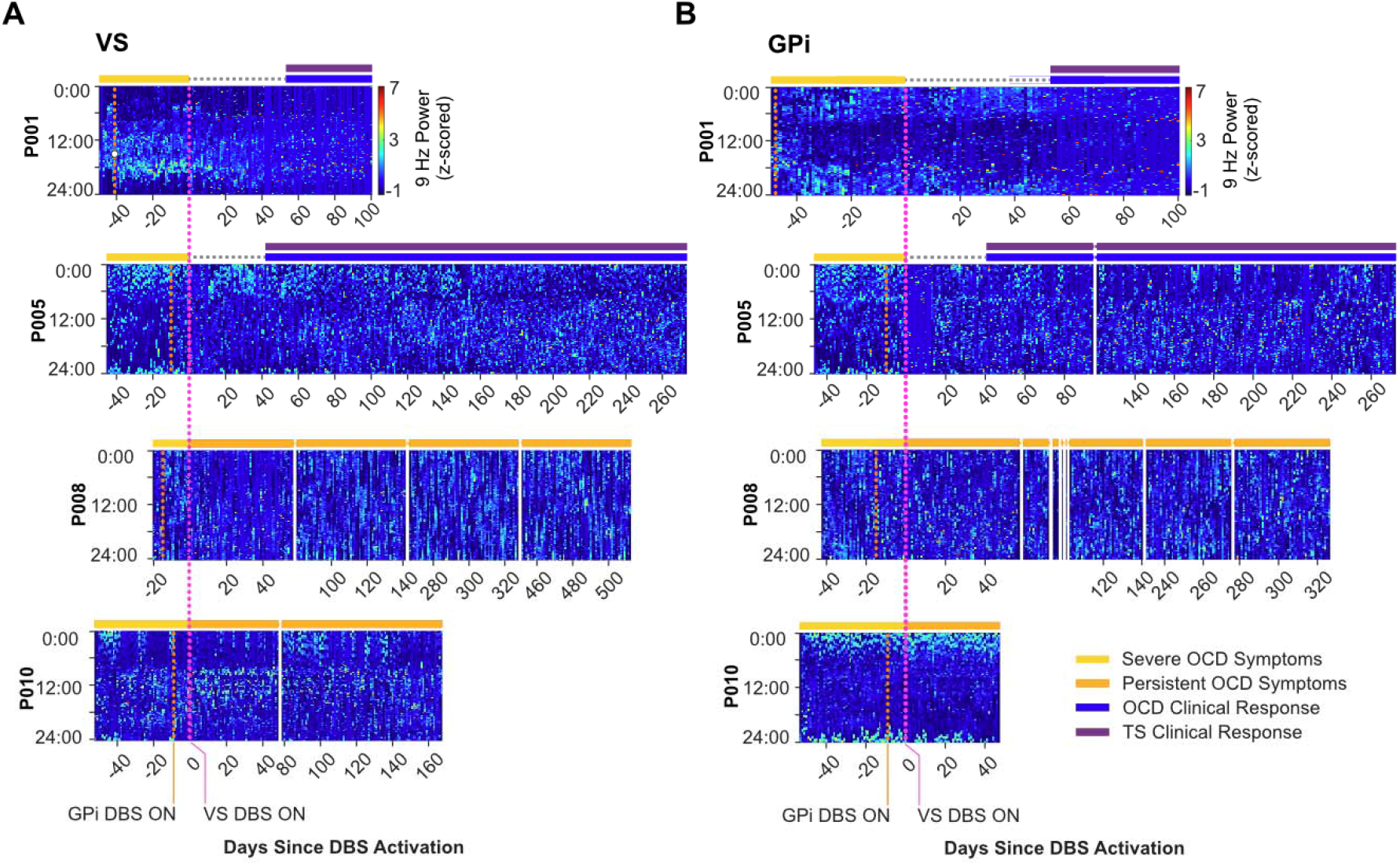
Circadian rhythmicity of 9 Hz spectral power in GPi and VS on and off chronic DBS. Heatmaps showing 9 Hz spectral power (z-score normalized per day) recorded from the a) GPi and b) VS targets in all patients across time. Each column within every patient image represents a 24-hour period aligned to days since VS stimulation activation. Times of GPi and VS stimulation onset are marked with vertical orange and magenta dashed lines, respectively. Both clinical responder patients (P001 and P005), exhibited sustained disruption of neural circadian rhythmicity during continuous stimulation that aligned with clinical response. In contrast, no disruption was evident in non-responder patients (P008 and P010).

**Figure 3.**
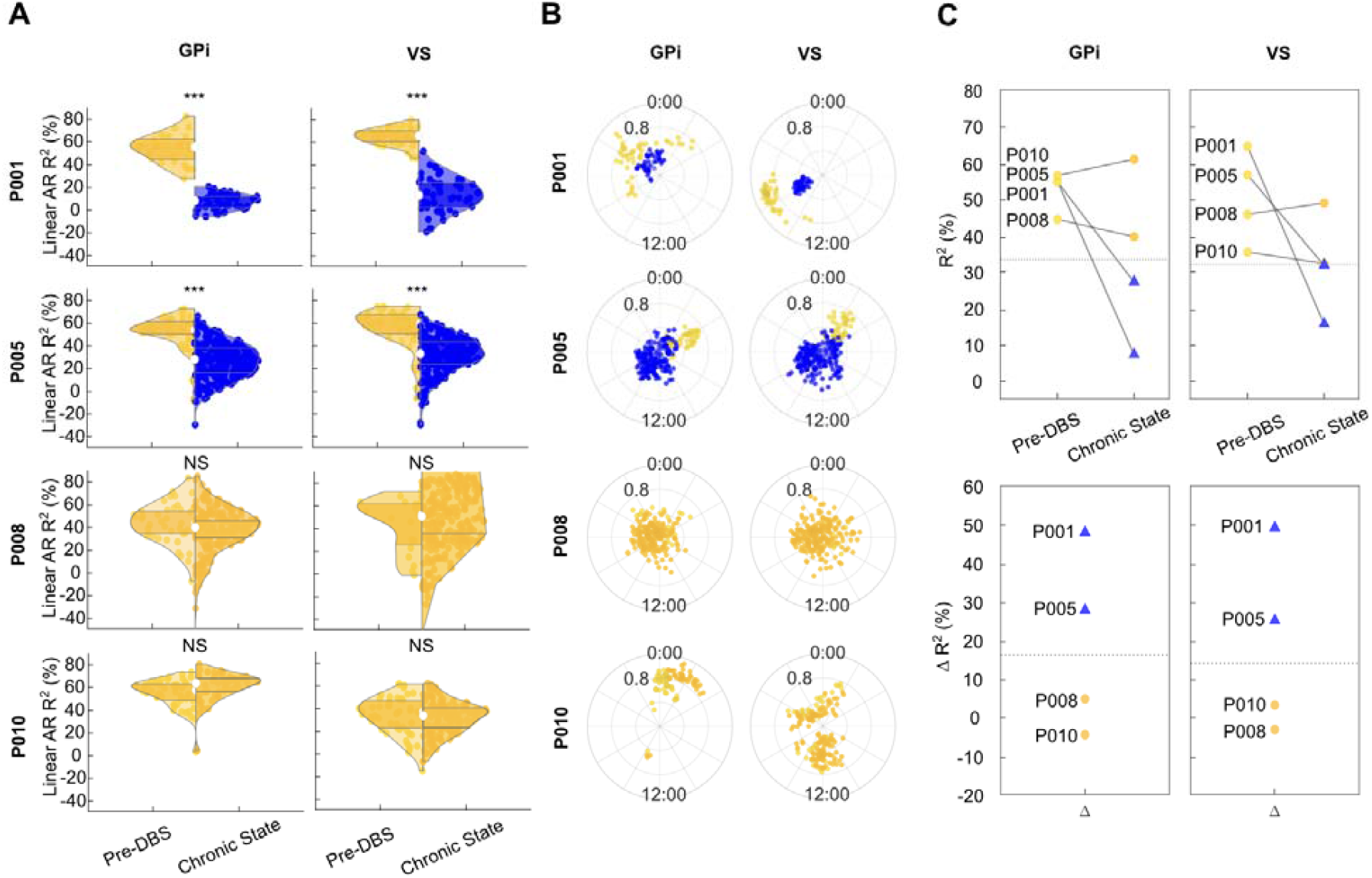
Disruption of linear predictability can classify responders and non-responders in the GPi and VS. a) Distribution of linear autoregressive (AR) model predictability using coefficient of determination (R^2^). Individual samples show the model prediction performances of the following day. A decrease in R^2^ during the chronic stimulation state is visible for stimulation responders in both GPi and VS targets. b) In addition to the AR model we fitted cosinor regressors that enabled investigation of circadian peak amplitude and phase changes with stimulation, which also showed neural periodicity disruption in responder patients. c) For each patient we fitted AR models and validated the mean R^2^ performance within five-fold cross-validation. The dashed lines show the maximum margin classifier between responder and non-responder patients. Responder and non-responder patients could thus be classified within both target regions by their respective neural circadian rhythmicity changes.

We found a significant reduction in the coefficient of determination (R^2^) in the chronic stimulation state in responders across both GPi (Linear AR R^2^ % P001 pre-stimulation 40.46 ± 24.08, post-stimulation 5.9 ± 7.17, Welch t-test p=10^−12^; P005 pre-stimulation 46.36 ± 19.48, post-stimulation 16.23 ± 11.54, Welch t-test p=10^−13^) and VS (Linear AR R^2^ P001 pre-stimulation 64.46 ± 7.47, post-stimulation 14.54 ± 12.62, Welch t-test p=10^−35^; P005 pre-stimulation 55.23 ± 18.4, post-stimulation 31.65 ± 16.71, Welch t-test p=10^−10^) (Fig. 3a). For the non-responder group, we observed in the GPi for one patient a non-significant reduction in neural predictability (Linear AR R^2^ % P008 pre-stimulation 43.72 ± 18.44, post-stimulation 34.83 ± 39.96, Welch t-test p=0.055) and notably in the other non-responder patient a significant increase (Linear AR R^2^ % P010 pre-stimulation 54.9 ± 12.55, post-stimulation 60.56 ± 12.68, Welch t-test p=0.027). Both non-responder patients exhibited in the VS non-significant neural predictability decreases (Linear AR R^2^ % P008 pre-stimulation 43.19 ± 23.95, post-stimulation 29.8 ± 280.86, Welch t-test p=0.48; P010 pre-stimulation 33.92 ± 17.14, post-stimulation 31.8 ± 14.6, Welch t-test p=0.44). We quantified with a cosinor regression model peak circadian phase and amplitude changes (Figure 3b). The peak circadian phases varied across patients and were modulated differently by stimulation. We reproduced notable changes in neurophysiological periodicity (Provenza et al., 2024): The linear AR model showed for responders a respective mean R^2^ reduction of 34% and 28% in GPi and 49% and 25% in VS. For non-responders the R^2^ reduction was absent for both patients: 4 and −4% in GPi and 3 and −3% in VS (Figure 3c).

### Correlations in activity across distant subcortical sites

Due to simultaneous dual-implant streamed recordings we could investigate the longitudinal circadian coupling characteristics between the two subcortical regions. First, we computed the direct correlations of 9 Hz power across GPi and VS sites. As a result of strong individual variability, we report the coupling characteristics separately for each patient. For patient P001 we found independent 9 Hz power fluctuations in each region, with an overall low correlation (Figure 4a, Pearson correlation coefficient r=0.02, p=0.002). For the other patients we found stronger correlation, though still with independent activity patterns (Pearson correlation coefficient P005 r=0.36 p<10^−5^, P008=0.22 p<10^−5^, P010=0.31 p<10^−5^).

**Figure 4.**
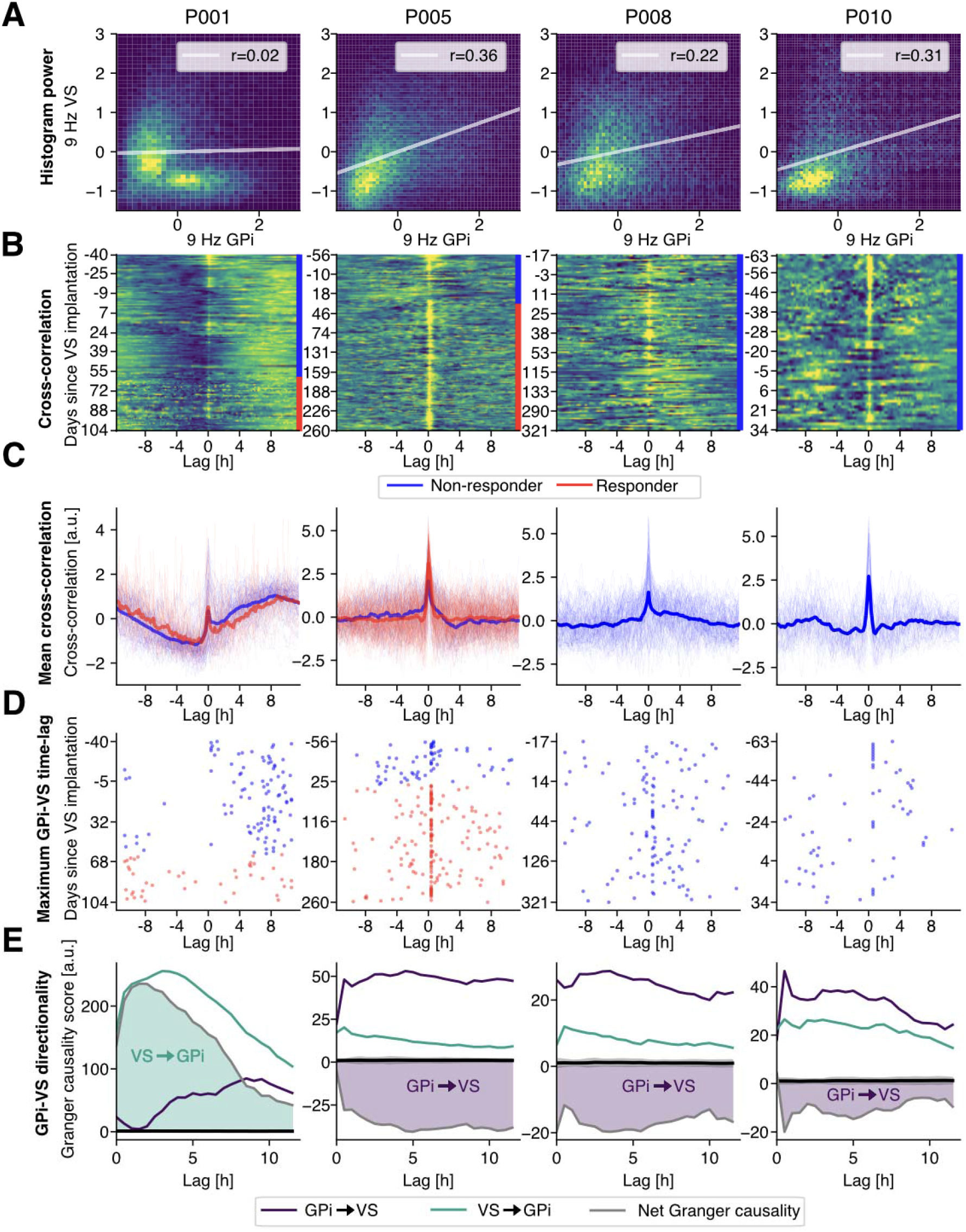
Simultaneous longitudinal 9 Hz GPi-VS recordings show patient-individual coupling characteristics. a) Joint GPi-VS 9 Hz power distributions show distinct instantaneous correlations (Pearson correlation coefficient) across four patients (P001, P005, P008, P010). P001 presents a bimodal distribution across the two sites. b) Daily cross-correlation patterns across days since VS stimulation. c) Individual cross-correlation traces color-coded for clinically effective OCD response. On average a strong zero-lag cross-correlation component is visible, overlayed with patient-individual independent coupling with different time-lags. d) Scatter plots show the maximum cross-correlation time-lag without zero-lag interaction. Mean time-lags deviated across patients, and did not show consistent coupling across responder and non-responder patients. Patients P001 presents strong sinusoidal coupling besides the zero-lag interaction. e) Granger-causality analysis showing 9 Hz power directionality between GPi and VS targets. Shaded areas indicate net directionality (GPi→VS minus VS→GPi). For P001 VS Granger-caused GPi 9 Hz power. For the other patients Granger-score directionality was reversed.

We then investigated inter-region coupling by computing cross-correlations of 9 Hz power between GPi and VS targets for each recording day (Figure 4b). For all patients we found a strong instantaneous zero-lag cross-correlation component that sharply decayed with increasing time-lags (Figure 4c). For patient P001 this component was overlayed by a sinusoidal modulation that had its mean peak lag at 8.82 h. For the other patients the peak lag component, without the instantaneous component, was at the first 10 min power sample, indicating predominantly synchronous activity between GPi and VS regions. Patient P005 showed no coupling between targets outside of the instantaneous component. Patient P008 showed a consistent decaying positive cross-correlation component succeeding the zero-lag component, and P010 represented phasic coupling preceding and succeeding the instantaneous correlation. In the two responder patients, daily cross-correlation patterns prior to response were significantly correlated with the daily response patterns after stimulation (Pearson correlation permutation test between mean cross-correlation patterns P001 r=0.91, *p*<10^−5^; P005 r=0.75, *p*<10^−5^). Additionally, we quantified the time-lag of the maximum cross-correlation pattern across days (excluding the zero-lag correlation component) (Figure 4d). Patient P001 exhibited significant modulation of peak time-lag between non-response and response periods (permutation test peak time-lag response vs non-response times: p<10^−5^), while for patient P005 no change was found (permutation test peak time-lag response vs non-response times: p=0.32).

The presence of a similar signal raises the question of its source. We examined two possibilities: one would be that one site is generating a signal that is then transmitted to the other. Alternatively, it could be that they are both inheriting this signal from a third source. To quantify causal directionality between regions, we computed Granger-causality scores within a 12 h time-lag window (Figure 4e). Here we estimated Granger causality in both directions and computed the net score by the individual score subtraction. Patient P001 exhibited strong directionality from VS to GPi, while for the remaining patients the directionality was reversed, with less strong overall scores. This finding suggests that across patients the 9 Hz power coupling can be driven in both directions and that it is likely that alternative sources are generating these signals.

### High-resolution time-domain streaming does not reveal a 9 Hz correlation

To investigate whether the coupling could also be identified in high-resolution temporal signals, we recorded high-resolution time-domain recordings in both targets in a single patient (patient P010) (Figure 5a). In comparison to the longitudinal recordings with a recorded power-snapshot sample every 10 min, these high-resolution time-domain recordings were acquired with a sampling rate of 250 Hz and synchronized via an external trigger as described previously (Provenza et al., 2021). We first computed time-resolved power in both targets (Figure 5b) and correlated the instantaneous power across targets. Contrary to the longitudinal reported positive 9 Hz power correlation (Figure 4a, patient P010), we did not detect the same strong positive correlation in the 9 Hz power or raw amplitude (Spearman correlation permutation test raw amplitude ρ = 0.03, p<10^−5^, 9 Hz power ρ = −0.02, p<10^−5^) (Figure 5c). To investigate frequency-specific synchronization within the high temporal-resolution recording, we computed coherence across the two sites. We identified strong coupling in the high beta to low-gamma range exceeding permuted chance level but absent synchronization in the 9 Hz band (Figure 5d).

**Figure 5:**
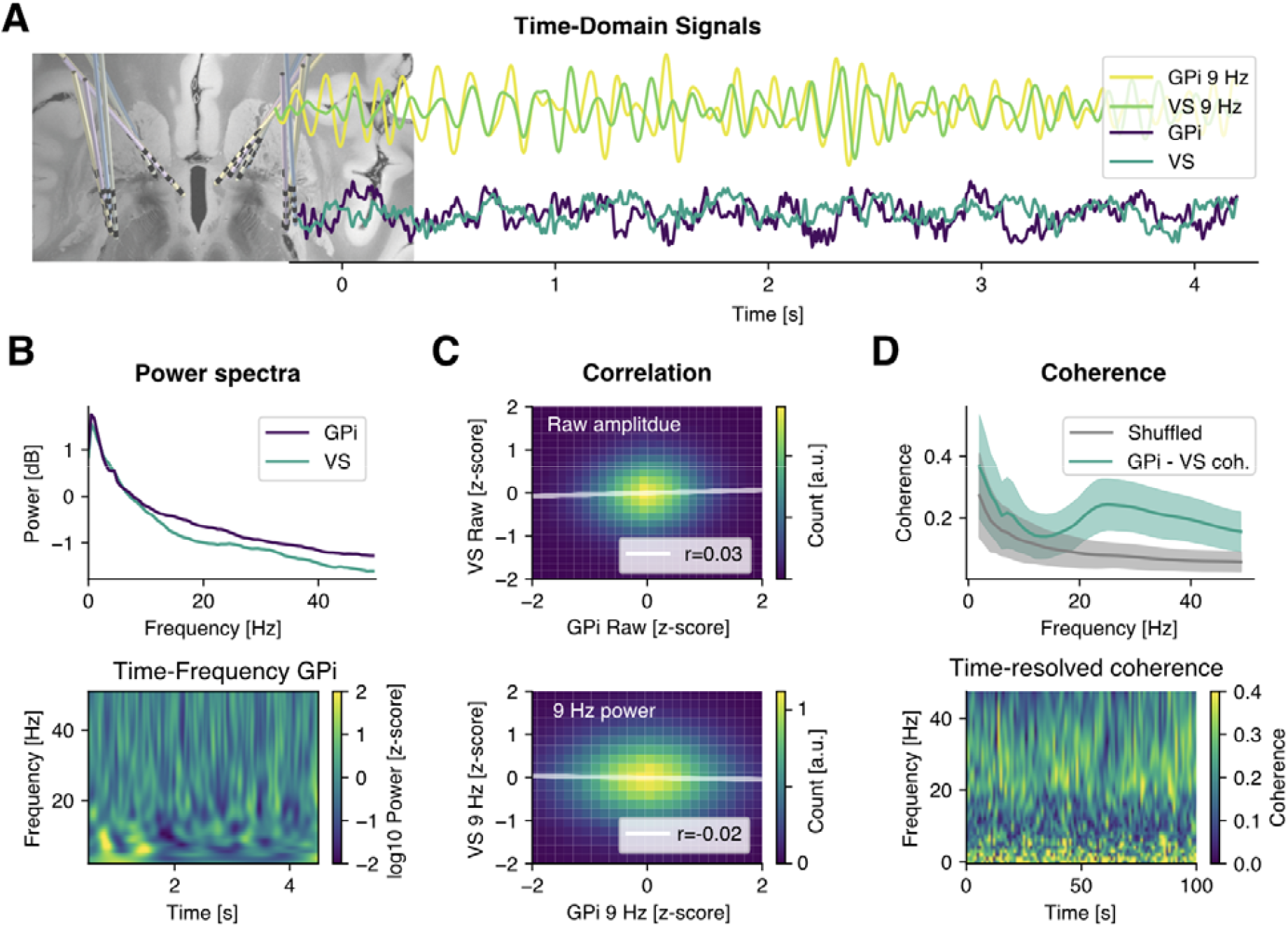
High-resolution streaming data reveals no raw signal or 9 Hz power correlation between GPi and VS targets. a) Time-domain traces from simultaneous GPi and VS recordings for both raw LFP and 9 Hz bandpass-filtered signals. b) Average power spectral density of GPi and VS LFP recordings (top), and time-frequency representation of GPi activity that shows transient bursts of 9 Hz oscillatory activity (bottom). c) Joint GPi-VS distribution histograms of normalized raw amplitudes and 9 Hz band power. Both distributions indicate low correlation. d) Magnitude-squared coherence between GPi and VS signals (green) shows strong coupling in the beta band exceeding time-shuffled control in gray. Shaded areas represent standard error across 10 s epochs. Time-resolved coherence spectrogram shows transient high beta (20-30 Hz) coupling between GPi and VS targets (bottom).

## Discussion

Our study presents evidence for the association between neural circadian periodicity modulation and clinically effective DBS in multiple brain regions. We showed previously that the disruption of this pattern could predict clinically effective stimulation in OCD patients in the VS (Provenza et al., 2024). Given the unique opportunity of dual-target DBS for comorbid OCD and TS in this cohort, we investigated the presence and coupling of neural circadian rhythmicity in an anatomically distinct brain region. In this new cohort, we found that the disrupted neural periodicity we originally observed in the VS after clinical response extends to another region in the basal ganglia, the GPi. These results indicate that this phenomenon is not unique to the VS and may be an indication of successful target engagement.

The overall effect size between changes in quantified disruption of neural circadian periodicity was similar between sites (Figure 3a) but showed clear differences in circadian phase preferences (Figure 3b). Previous research in a large cohort of Parkinson’s disease DBS patients demonstrated circadian phase preferences in STN or GPi that were heterogeneous within each target across patients (Cagle et al., 2024). Our results underscore this heterogeneity, which we now also show in a multi-target approach in psychiatric patients.

This dual-target DBS cohort also allowed us to investigate coupling between circadian neural periodicity in multiple DBS target regions within the same patients. Here we found strong variability in longitudinal 9 Hz power coupling across patients. Neither responders nor non-responders exhibited consistent instantaneous power correlation, circadian cross-correlation patterns or net Granger causality (Figure 4). In responders, we observed significant cross-correlation between VS and GPi, even in the presence of the overall disruption of neural predictability. Further investigation is needed to uncover the mechanisms behind inter-patient variability in neural periodicity. Patient-individual anatomical differences might be contributing factors (Wen et al., 2025) in addition to variations in comorbid conditions and subcortical lead placements.

The neural circuits underlying the presented neural rhythmicity specifically in the VS and GPi target structures require further exploration. The VS is the limbic component of the striatum, receiving inputs from reward and motivation-related cortical structures (Haber et al., 2006). It projects to the ventral pallidum, which has components that are similar to both the GPe (indirect pathway) and GPi (direct pathway) (Heimer, 1972; Heimer et al., 1982; Nauta et al., 1978; Root et al., 2015). However, pallidal projections from the VS are not limited to the ventral compartment; they extend to medial, anterior, and ventral portions of the pallidum, including the GPi (Haber et al., 1990; Hazrati and Parent, 1992; Heilbronner et al., 2018). The GPi target in this case is quite posteriorly situated relative to published maps of VS projections. Still, we cannot eliminate the possibility of a (likely modest) direct projection from VS to GPi (while the GPi does not project to the striatum). Finally, although these targets are named “VS” and “GPi,” the VTAs likely encompass neighboring structures, such as the ventral anterior limb of the internal capsule and the GPe, respectively. However, these additional structures are likely not a source of shared axons/cell bodies. More likely is that there is a consistent pattern across many structures, or that there is shared input allowing the VS and GPi to coordinate. Along these lines, there is strong evidence that neural circadian patterns are generated in the suprachiasmatic nucleus (SCN) of the hypothalamus. The SCN does not project directly to the GPi or VS, but multisynaptic relays may allow it to influence large portions of the brain (Patton and Hastings, 2018).

### Opportunities for closed-loop neuromodulation

Insights into the relationship between neural signals and symptom states will inform strategies that use these signals either to assess target engagement and/or as feedback in closed loop implementations. (Fleming et al., 2022; Toth et al., 2020). Here we show that in addition to neural circadian rhythmicity in the stimulation target region, there can be another distinct subcortical region, that could be used as a source for control sensing signals. This dual-target approach could be advantageous when the local biomarker signal might be contaminated by stimulation artifacts. In addition, a combination of different temporal feedback signals might be integrated to account for short- and long-term symptoms. We showed that the longitudinal power fluctuations in the 9 Hz band were not present during short-term high-resolution temporal recordings in one exemplary patient (Figure 5). Since OCD symptoms manifest on multiple timescales both as short-term episodes of acute distress and long-term patterns of maladaptive behavior and cognitive rigidity, different adaptive DBS strategies could be tailored to target transient symptom exacerbations as well as sustained pathological network activity (Figee et al., 2013; Provenza et al., 2024, 2021).

In summary, we investigated circadian rhythmicity in subcortical neural activity using chronic dual-site recordings from patients with comorbid OCD and TS treated with bilateral DBS in the VS and GPi. Using continuous longitudinal sensing, we show that neural periodicity in the 9□Hz band was disrupted following clinically effective stimulation in both regions. Time-resolved spectral analysis and autoregressive modeling revealed that this disruption was predictive of therapeutic response and absent in non-responders in both regions. Inter-regional analyses showed that coupling between GPi and VS circadian dynamics was patient-specific, with variable delays and directionality. We show that there can be independent neural circadian patterns in VS and GPi that are both predictive of clinical DBS response. Denser behavioral measures would enable higher resolution investigation of these neurobehavioral relationships and are currently underway. Overall, our findings characterize multi-site subcortical neural chronobiology in psychiatric and movement disorders and highlight circadian biomarkers as promising signals for assessing target engagement and possibly for guiding future closed-loop DBS strategies.

## Data Availability

All data produced in the present study are available upon reasonable request to the authors

## Disclosures

Dr. Storch reports receiving research funding to his institution from the Ream Foundation, International OCD Foundation, and NIH. He was a consultant for Brainsway and Biohaven Pharmaceuticals in the past three years. He owns stock less than $5000 in NView/Proem for distribution of the YBOCS Scales. He receives book royalties from Elsevier, Wiley, Oxford, American Psychological Association, Guildford, Springer, Routledge, and Jessica Kingsley.

Dr. W. K. Goodman receives royalties from Nview, LLC and OCDscales, LLC, and research support from NIH (UH3NS100549), IOCDF, and McNair Foundation.

## Notes

### Clinical Trial

NCT05915741

### Funding Statement

This research was supported by the National Institutes of Health (NIH) National Institute of Neurological Disorders and Stroke BRAIN Initiative via contract UH3NS136631 (to S.A.S., W.K.G., J.A.H., N.R.P.), the McNair Foundation (S.A.S., N.R.P.) and the Gordon and Mary Cain Pediatric Neurology Research Foundation (S.A.S.).

### Author Declarations

The study protocol was approved by the Institutional Review Board (IRB) at Baylor College of Medicine (H-49155)

